# Association of demographic factors with acute mountain sickness: a retrospective study in Litang

**DOI:** 10.1101/2024.06.18.24309071

**Authors:** Haitong Zhao, Hao Wang, Chao Wang, Lifen Li, Xuan Zhang, Ling Chen, Guohua Ni, Hui Yan, Lei Chen, Fengming Luo

**Author notes:** Correspondence: Fengming Luo MD and Lei Chen MD, Department of High Altitude Medicine, Center for High Altitude Medicine, West China Hospital, Sichuan University, Chengdu, Sichuan 610041, China. Contributed equally (Haitong Zhao, Hao Wang, Chao Wang).

## Abstract

**Objective:** Acute mountain sickness (AMS) is a common clinical syndrome in high altitude areas, but the role of demographic factors in AMS has not been well-illucidated. This study aims to describe the relationship between demographic factors and AMS in Litang, one of the highest-altitude town in the world.

**Methods:** Demographic data of patients diagnosed with AMS from January 2022 to December 2023 were collected from the information management system of Litang County People’s Hospital, including gender, age, onset time, ethnicity, current residence and its altitude. Descriptive statistical methods were used to describe these factors.

**Results:** 7,290 AMS patients were included, among which 62.37% (4,547/7,290) were male and 37.63% (2,743/7,290) were female; 90.41% (6,591/7,290) were non-Tibetan, and 9.59% (699/7,290) were Tibetan; the age group 20-29 had the highest number of patients (2,422, 39.08%), and the incidence of AMS generally decreased with age; the proportion of AMS occurring from May to August reached 51.99% (3,790/7,290); the percentage from lower altitude areas was 79.05% (4,333/7,150).

**Conclusions:** These data from Litang suggest higher incidence of AMS may be detected in males, non-Tibetans, young people, summer time and residents from lower altitude. Special attention should be paid to these demographic factors for preventing the occurrence of AMS.

## 1 Introduction

Acute Mountain Sickness (AMS) refers to a series of clinical syndromes that occur when individuals rapidly ascend from low altitude areas (≤1,500m) to high altitude areas (>2,500m), due to inadequate or dysregulated adaptation to the hypobaric hypoxic environment. The main manifestations of AMS are headaches, dizziness, nausea, vomiting, fatigue, and other discomforts. In severe cases, it can lead to life-threatening conditions such as high-altitude pulmonary edema and high-altitude cerebral edema^1-3^. The harsh, cold, hypoxic, dry conditions, and strong ultraviolet radiation in high-altitude areas are environmental factors that trigger stress responses such as increased cardiac output, which in turn cause AMS^4-6^.

Litang County, a new tourist destination, also called “City in the Sky”, is located in the western part of Sichuan province of China, and on the southeastern edge of the Tibetan Plateau, with an average altitude of 4,014 meters, making it one of the most highest-altitude town in the world. The geographic location of Litang County resulted in a hypobaric hypoxic high-altitude environment, with oxygen content in the air being only 48% of that in plain areas^7^. Additionally, the region experiences low temperatures, long winter, abundant sunlight, intensive radiation, and strong winds^8^. This harsh hypobaric hypoxic environment may trigger various high-altitude reactions, among which AMS is a common disease^9^.

Therefore, this study retrospectively analyzed the demographic data of AMS patients treated in Litang County People’s Hospital, and described the relationship between demographic factors and AMS, in order to prevent the occurrence of AMS.

## 2 Methods

### 2.1 Study population

All the AMS patients treated in Litang County People’s Hospital were recruited from January 2022 to December 2023, whose demographic data, including gender, age, time of onset, ethnicity, current residence and its altitude, were collected from the hospital information management system. These patients must meet the diagnosis criteria of AMS based on the Lake Louise Scoring: (1) headache; (2) at least one additional symptom (gastrointestinal symptoms, fatigue and/or weakness, dizziness/light-headedness); (3) the total score was equal to or more than 3^10,11^. Patients were excluded if they were aged <10 years old, or had history of mental diseases. The study was approved by the Ethics Committee of West China Hospital of Sichuan University (No. 2023-1164).

### 2.2 Statistical analysis

Continuous data were expressed as means and standard deviations, while binary data were expressed as percentages. Excel 2019 was used for statistical analysis and graphing.

## 3 Results

### 3.1 Gender, ethnicity and age

7,290 AMS patients were included, among which 4,547 were male, accounting for 62.37%, and 2,743 were female, accounting for 37.63%; non-Tibetans accounted for 6,591 (90.41%), while Tibetans comprised 699 (9.59%).

Excluding 245 patients without age data, the average age of AMS patients was 34.98 years old, with the average age for males being 34.91 and females 35.05. In the population aged 20 and above, the largest number of patients fell within the 20-29 age group (2,422, 39.08%), followed by those aged 30-39 (2,202, 31.26%). The incidence of AMS generally showed a decreasing trend with increasing age (Figure 1).

**Figure 1:**
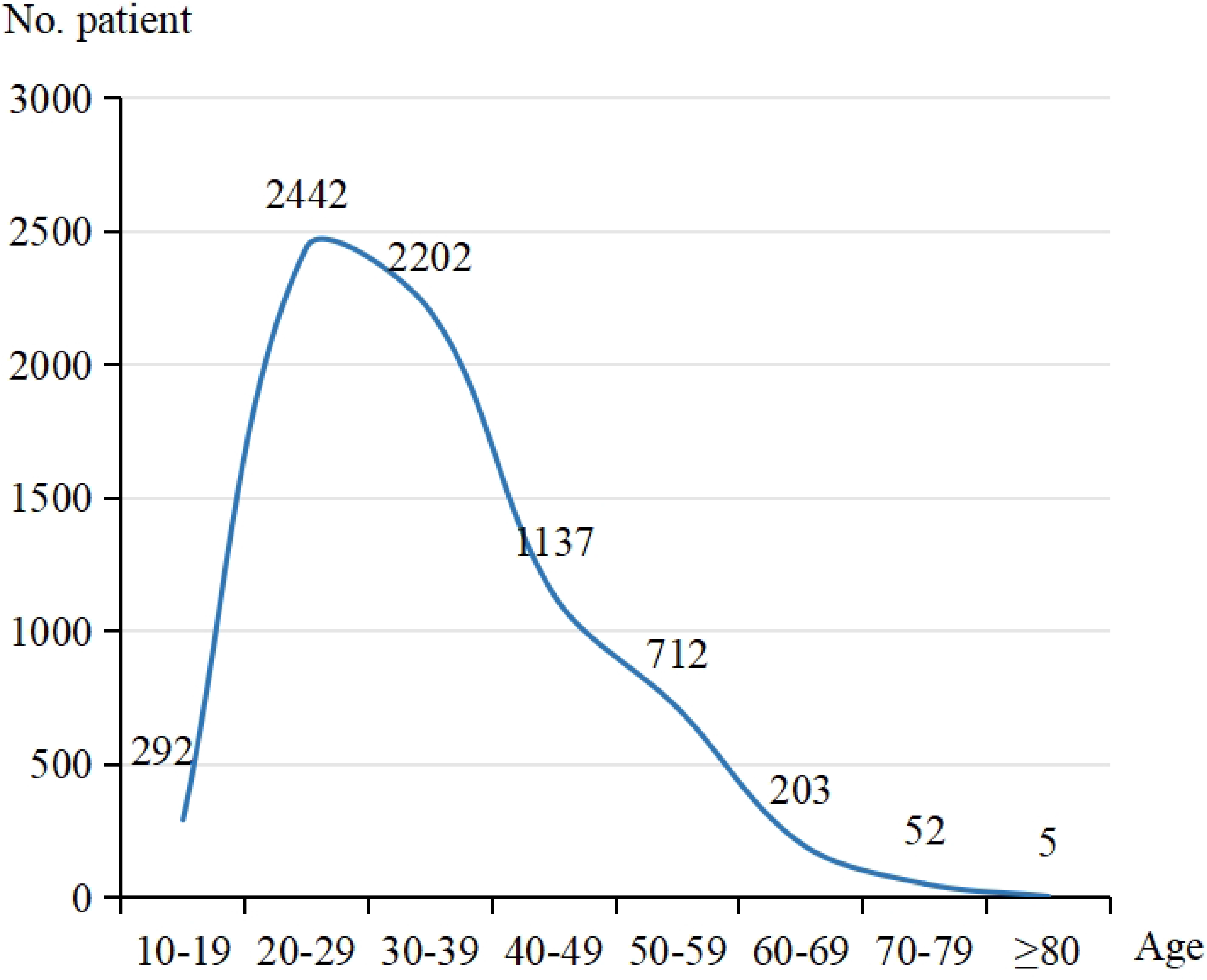
Distribution of age for AMS patients in Litang.

### 3.2 Time of onset

In July, Litang County People’s Hospital treated the highest number of AMS patients (1,456, 19.97%), followed by May (842, 11.55%), August (781, 10.71%), and June (711, 9.75%) (Figure 2). The number of cases during these four months accounted for 51.99% of the total patients.

**Figure 2:**
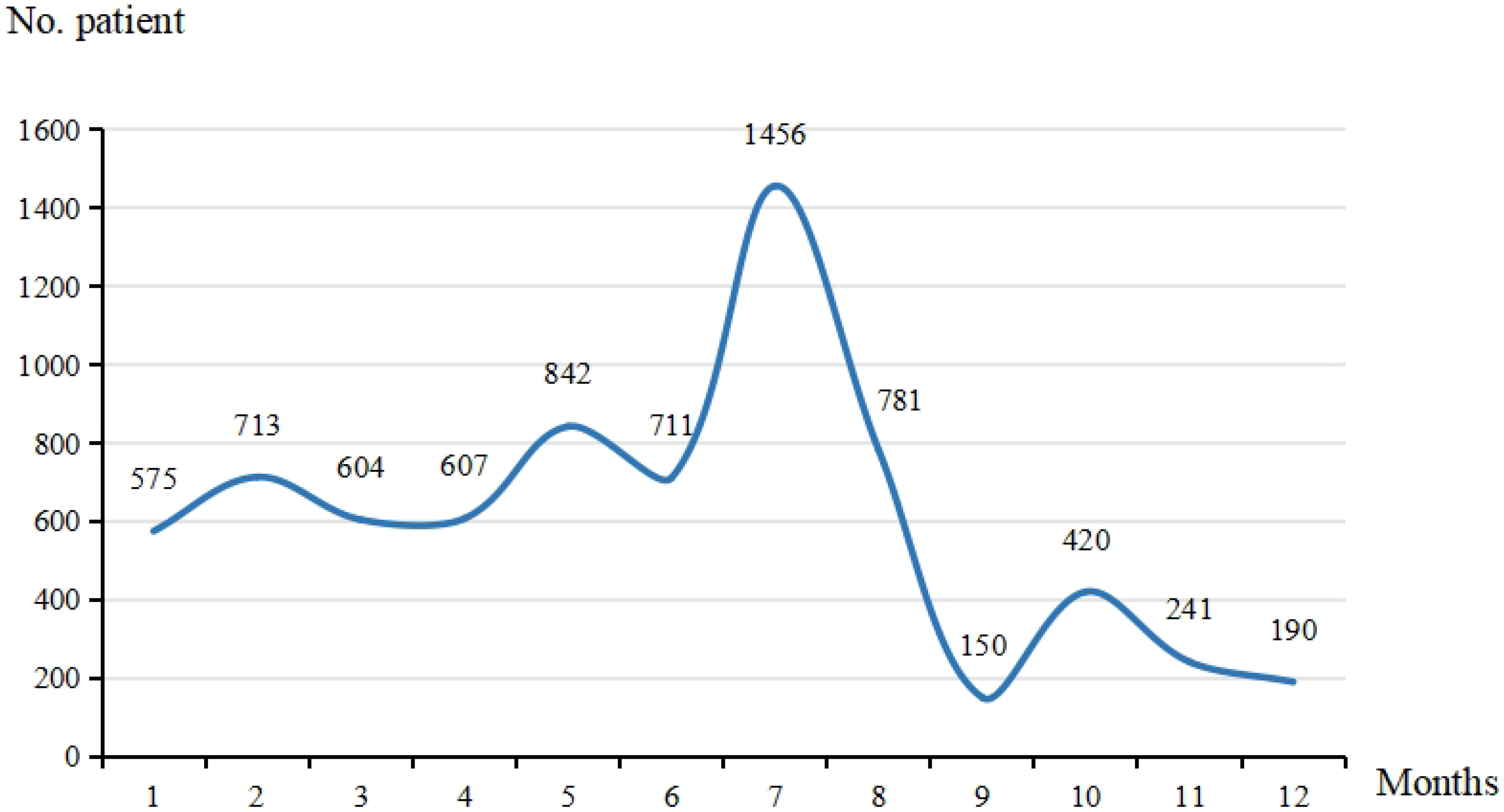
Distribution of time of onset for AMS patients in Litang.

### 3.3 Residental altitude

Excluding 140 AMS patients without data reagrding current residence and its altitude, 2,817 (39.40%) came from Sichuan Province, and 4,333 (60.60%) came from outside Sichuan Province. Among these AMS patients, 79.05% came from low-altitude areas, with 32.84% residing at altitudes of 0-100 meters and 30.55% at 101-500 meters; 20.95% came from high-altitude areas (Figure 3). Moreover, analysis of 4,624 AMS patients aged 20-29 years and 30-39 years showed that 78.53% came from low-altitude areas and 21.47% came from high-altitude areas.

**Figure 3:**
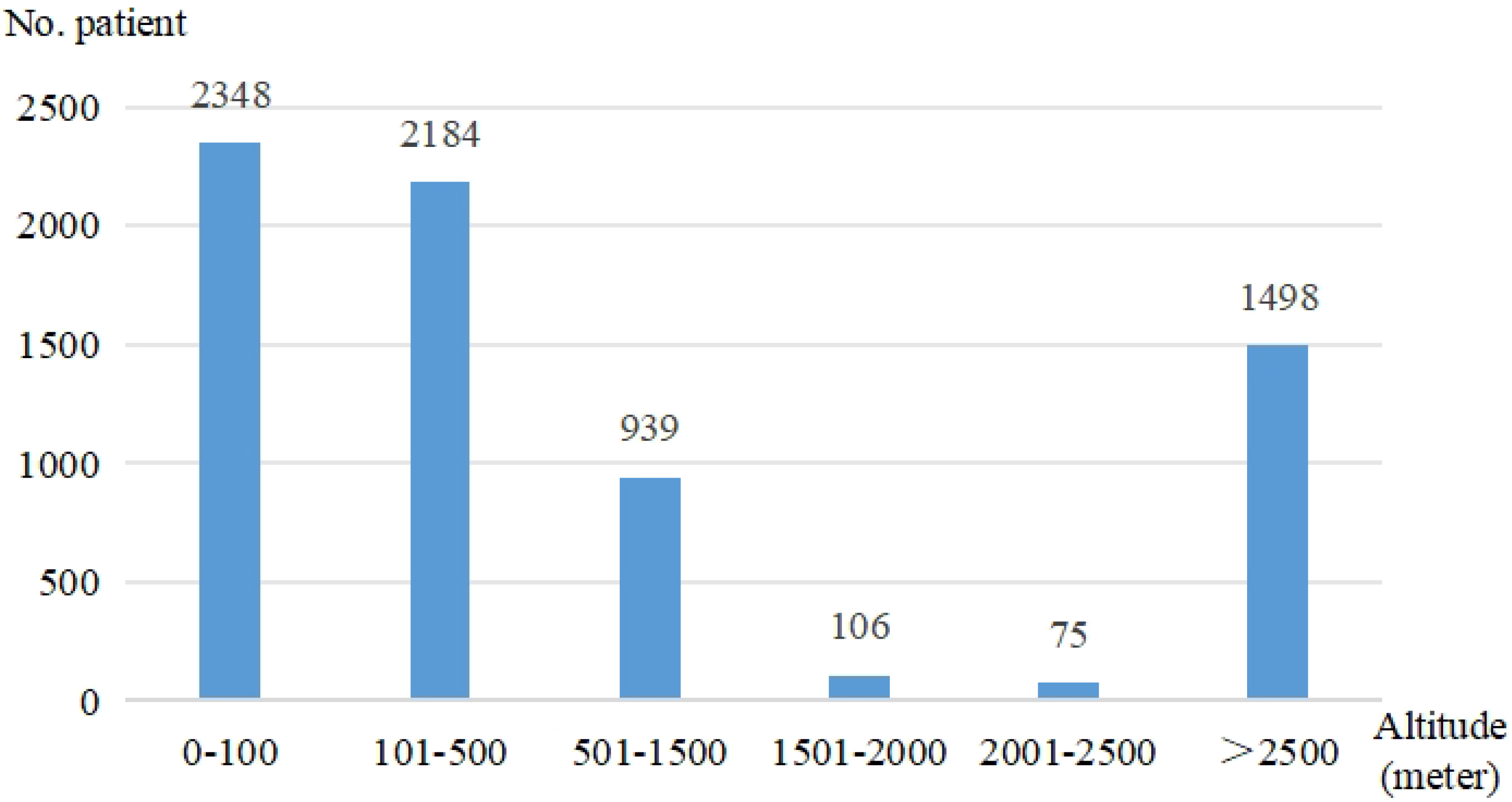
Distribution of residental altitude for AMS patients in Litang.

## 4 Discussion

This study showed that among the AMS patients at Litang County People’s Hospital from January 2022 to December 2023, there were higher proportions in males, non-Tibetans, individuals aged 20-39, those visiting during May to August, and those from low-altitude areas.

Sex-based differences in the prevalence of acute mountain sickness was discussed recently, which informed higher prevalence rate of AMS in women than in men, however, the absolute number of AMS in men was much more than that in women^7^. Similarly, our data showed that males accounted for a greater proportion of AMS patients.

Tibetans have lived in high-altitude regions for tens of thousands of years. Therefore, relative to non-Tibetans, their bodies have a clear genetic basis for adaptation to high-altitude conditions, enabling them to quickly adjust to low-oxygen environments and making them less susceptible to AMS and other altitude-related illnesses^12-14^. which was definitely indicated in the present study.

In recent years, Litang County has been enhancing its cultural and tourism capabilities, which is attracting a significant number of visitors, particularly young visitors from the country. As a result, in this study, young peolpe aged 20-39 had the highest proportion of AMS. Moreover, according to the Litang County Tourism Bureau, summer time is the peak tourist season. Consequencely, this study revealed the highest number of AMS cases from May to August, which was overlapped with the peak tourist season.

In this study, we found people from lower altitude of resident place had the higher proportions of AMS occurence, which indicated migrants from lower altitude areas were much more prone to AMS, mainly owing to unacclimatization to the hypobaric hypoxic environment of high altitudes^1,15^. Moreover, air travel has become the preferred way of transportation for most tourists, but a large observational study found that when tourists rapidly ascended to high-altitude destinations by air, the incidence of AMS increased by 4.5 times; in contrast, the incidence increased only by 2.1% for those who travelled by foot or a combination of walking, driving and flying^2^. Therefore, a gradual ascent strategy should be adopted, staying at different altitude gradients to prolong exposure to high altitudes, thereby reducing the incidence of AMS, and preventing more severe conditions such as high-altitude pulmonary and cerebral edema^16,17^.

Noticeably, a high proportion of AMS was also revealed in high altitude population, demonstrating residents in high altitude areas, especially the migrants, still confront the risk of AMS, because of unacclimatization (inadaptation) to the hypobaric hypoxic environment, and other factors, including acute respiratory tract infections, strenuous activities, etc^3,18,19^.

## 5 Conclusions

This retrospective study suggests, based on the data from Litang, higher incidence of AMS may be detected in males, non-Tibetans, young people, summer time and residents from lower altitude. Special attention should be paid to these demographic factors for preventing the occurrence of AMS.

## Data Availability

All relevant data are within the manuscript and its Supporting Information files

## References

1. Prince TS, Thurman J, Huebner K. Acute Mountain Sickness. In: StatPearls. Treasure Island (FL): StatPearls Publishing Copyright © 2024, StatPearls Publishing LLC.; 2024.

2. Burtscher J, Swenson ER, Hackett PH, Millet GP, Burtscher M. Flying to high-altitude destinations: Is the risk of acute mountain sickness greater? J Travel Med. 2023;30(4) doi: 10.1093/jtm/taad011

3. Zubieta-Calleja GR, Zubieta-DeUrioste N. High Altitude Pulmonary Edema, High Altitude Cerebral Edema, and Acute Mountain Sickness: an enhanced opinion from the High Andes - La Paz, Bolivia 3,500 m. Rev Environ Health. 2023;38(2):327–338. doi: 10.1515/reveh-2021-0172

4. Bian SZ, Jin J, Zhang JH, Li QN, Yu J, Yu SY, Chen JF, Yu XJ, et al. Principal Component Analysis and Risk Factors for Acute Mountain Sickness upon Acute Exposure at 3700 m. PLoS One. 2015;10(11):e0142375. doi: 10.1371/journal.pone.0142375

5. DeLellis SM, Anderson SE, Lynch JH, Kratz K. Acute mountain sickness prophylaxis: a high-altitude perspective. Curr Sports Med Rep. 2013;12(2):110–114. doi: 10.1249/JSR.0b013e3182874d0f

6. Swenson ER. Carbonic anhydrase inhibitors and high altitude illnesses. Subcell Biochem. 2014;75:361–386. doi: 10.1007/978-94-007-7359-2_18

7. He B, Feng J, Shu Y, Yang L, He Z, Liao K, Zhuo H, Li H. Prevalence and Risk Factors of Hyperuricemia Among Young and Middle-Aged Tibetan Men Living at Ultrahigh Altitudes: A Cross-Sectional Study. High Alt Med Biol. 2024;25(1):42–48. doi: 10.1089/ham.2023.0056

8. Luo Y, Yang X, Gao Y. Strategies for the prevention of acute mountain sickness and treatment for large groups making a rapid ascent in China. Int J Cardiol. 2013;169(2):97–100. doi: 10.1016/j.ijcard.2013.08.059

9. Huang P, Ke G, Lin X, Wang Q, Lu W, Zeng L, Xu S. Correlation analysis between vitamin A, D, and E status with altitude, seasonal variation, and other factors, among children aged 0-6 years in a Chinese population living in the Tibetan plateau of Ganzi prefecture. J Clin Lab Anal. 2022;36(9):e24620. doi: 10.1002/jcla.24620

10. Roach RC, Hackett PH, Oelz O, Bärtsch P, Luks AM, MacInnis MJ, Baillie JK. The 2018 Lake Louise Acute Mountain Sickness Score. High Alt Med Biol. 2018;19(1):4–6. doi: 10.1089/ham.2017.0164

11. Woolcott OO. The Lake Louise Acute Mountain Sickness Score: Still a Headache. High Alt Med Biol. 2021;22(4):351–352. doi: 10.1089/ham.2021.0110

12. Hou YP, Wu JL, Tan C, Chen Y, Guo R, Luo YJ. Sex-based differences in the prevalence of acute mountain sickness: a meta-analysis. Mil Med Res. 2019;6(1):38. doi: 10.1186/s40779-019-0228-3

13. Peng Y, Yang Z, Zhang H, Cui C, Qi X, Luo X, Tao X, Wu T, et al. Genetic variations in Tibetan populations and high-altitude adaptation at the Himalayas. Mol Biol Evol. 2011;28(2):1075–1081. doi: 10.1093/molbev/msq290

14. Chen B, Li D, Ran B, Zhang P, Wang T. Key miRNAs and Genes in the High-Altitude Adaptation of Tibetan Chickens. Front Vet Sci. 2022;9:911685. doi: 10.3389/fvets.2022.911685

15. Liu B, Xu G, Sun B, Wu G, Chen J, Gao Y. Clinical and biochemical indices of people with high-altitude experience linked to acute mountain sickness. Travel Med Infect Dis. 2023;51:102506. doi: 10.1016/j.tmaid.2022.102506

16. Roach RC, Maes D, Sandoval D, Robergs RA, Icenogle M, Hinghofer-Szalkay H, Lium D, Loeppky JA. Exercise exacerbates acute mountain sickness at simulated high altitude. J Appl Physiol (1985). 2000;88(2):581–585. doi: 10.1152/jappl.2000.88.2.581

17. Hsu TY, Weng YM, Chiu YH, Li WC, Chen PY, Wang SH, Huang KF, Kao WF, et al. Rate of ascent and acute mountain sickness at high altitude. Clin J Sport Med. 2015;25(2):95–104. doi: 10.1097/jsm.0000000000000098

18. Cobb AB, Levett DZH, Mitchell K, Aveling W, Hurlbut D, Gilbert-Kawai E, Hennis PJ, Mythen MG, et al. Physiological responses during ascent to high altitude and the incidence of acute mountain sickness. Physiol Rep. 2021;9(7):e14809. doi: 10.14814/phy2.14809

19. Gonggalanzi, Labasangzhu, Nafstad P, Stigum H, Wu T, Haldorsen Ø D, Ommundsen K, Bjertness E. Acute mountain sickness among tourists visiting the high-altitude city of Lhasa at 3658 m above sea level: a cross-sectional study. Arch Public Health. 2016;74:23. doi: 10.1186/s13690-016-0134-z

